# Plasma proteome demonstrates sex-specific associations with mental health risks in adolescents

**DOI:** 10.1101/2024.12.16.24319062

**Authors:** Alexey M. Afonin, Aino-Kaisa Piironen, Jordi Julvez, Irene van Kamp, Katja M. Kanninen

**Affiliations:** A.I. Virtanen Institute for Molecular Sciences, University of Eastern Finland, Kuopio, Finland; Clinical and Epidemiological Neuroscience (NeuroÈpia), Institut d’Investigació Sanitària Pere Virgili (IISPV), 43204 Reus, Spain; ISGlobal, Barcelona, Spain; CIBER Epidemiología y Salud Pública (CIBEREsp), Madrid, Spain; Centre for Sustainability, Environment and Health, National Institute for Public Health and the Environment, Bilthoven, the Netherlands

## Abstract

Adolescence is a critical developmental period marked by significant physiological, psychological, and behavioural changes. Sex-specific biological factors can play a major role in their progression. Liquid chromatography – tandem mass spectrometry proteomic analysis was used to measure the plasma proteome abundances in 197 adolescents (11-16 years old) from the WALNUTs cohort. Orthogonal partial least squares discriminant analysis (OPLS-DA) revealed clear sex-based proteomic distinctions, with 76 proteins significantly differing between males and females after correcting for age and BMI. Gene Ontology enrichment analysis of these proteins highlighted pathways related to cell adhesion and extracellular matrix organization reflecting sex-specific developmental trajectories during puberty. Bioinformatic analysis revealed 37 proteins significantly associated with the total score of the Strengths and Difficulties Questionnaire (SDQ), with additional sex-specific associations emerging in subgroup analyses. Plasma protein abundancies in males exhibited stronger correlations with SDQ externalizing subscale scores, while in females the associations with the internalizing score were more prominent, consistent with known behavioural sex differences. Immune response and blood coagulation pathways were implicated in these associations, particularly in females, while no significant pathway enrichment was observed for males. These findings highlight both shared and sex-specific proteomic features associated with the SDQ scores in adolescents, emphasising the need to consider sex differences in proteomic studies. The results provide a critical step toward identifying biomarkers and pathways underlying sex-specific psychological and developmental processes in adolescence.

## Introduction

Adolescence is a critical developmental period characterised by profound physiological, hormonal, and metabolic changes, many of which are sex specific. These changes have long-term implications for health and disease trajectories, making it a crucial time to explore biological differences between females and males. As is the case with many diseases, sex also plays a role in mental health. For instance, depression and anxiety disorders are among the most prevalent mental health conditions worldwide, with significant sex differences observed in their prevalence, manifestation, and treatment responses[1]. Epidemiological studies consistently report that women can be more than twice as likely as men to experience depression and anxiety disorders [2–4]. Men, while exhibiting lower reported rates of these disorders, may underreport symptoms due to social stigma and are more likely to suffer from other problems, such as substance abuse[1].

The Strengths and Difficulties Questionnaire (SDQ) is a widely used screening tool assessing behavioural and emotional difficulties in children and adolescents across five domains: emotional symptoms, conduct problems, hyperactivity/inattention, peer relationship problems, and prosocial behaviour [5–7]. The psychological measures such as the SDQ and the general psychopathology factor (p-factor) can both be influenced by the sex of the participant. Understanding these differences is crucial for accurate diagnosis, intervention, and support across developmental stages [8]. Boys tend to exhibit higher scores in hyperactivity/inattention and conduct problems, reflecting a greater prevalence of externalizing behaviours [9, 10]. In contrast, girls often score higher in emotional symptoms and peer relationship problems, indicating a tendency toward internalizing issues [11]. These differences may be attributed to biological factors, socialization processes, and gender norms influencing the expression and reporting of psychological symptoms [12]. Sex-associated differences in mental disorders were reported in multiple papers using different OMIC technologies [13–16].

The plasma proteome can provide important insights into health and disease, but also into differences between the sexes. Several studies have identified plasma proteins with significantly different levels between adult females and males[17–19]. For instance, the apolipoproteins, integral to lipid transport and metabolism, often display sex-specific variations. Higher levels of apolipoprotein A-I have been observed in females, potentially offering protection against cardiovascular diseases[17]. Conversely, males may exhibit higher levels of proteins associated with inflammatory responses, such as C-reactive protein (CRP), a known risk marker for cardiovascular events [20]. A study of circulating cardiometabolic-related proteins associated with the risk of incident myocardial infarction revealed significant sex-specific differences. Among over 11,000 Swedish adults, 45 proteins were linked to MI risk, with 13 showing sex-specific associations—most prominently among women [21]. The immune system generally exhibits notable sex-specific characteristics: females generally mount stronger innate and adaptive immune responses than males, which is thought to contribute to the increased prevalence of autoimmune diseases in women[22]. Proteins such as interferons, interleukins, and immunoglobulins may be differentially expressed, influencing immune cell function and signalling pathways [23].

Sex-specific differences in mental health have been increasingly revealed through proteomics and other omics studies, highlighting distinct molecular mechanisms underlying psychiatric disorders between males and females. For instance, an untargeted proteomic analysis identified sex-specific biological abnormalities in late-life depression, finding 33 proteins uniquely associated with depression in females—primarily related to immunoinflammatory control—and 6 unique proteins in males, affecting a broader range of biological pathways [24]. Similarly, a metabolomics study in children and adolescents with major depressive disorder identified sex-specific plasma biomarkers, with biliverdin as a male-specific biomarker and phosphatidylcholine as a female-specific biomarker [14]. Genomic studies further support these findings; for example, Labonté et al. discovered sex-specific transcriptional signatures in depression, demonstrating different gene expression patterns in males and females [25]. These insights underscore the importance of considering sex as a critical variable in mental health research, with significant implications for developing personalised diagnostic tools and therapeutic interventions tailored to sex-specific biological profiles.

Most studies thus far have concentrated on assessing sex-specific plasma protein levels in adults, and relatively little is known about the situation in adolescents. A longitudinal study of plasma proteome of children and adolescents showed multiple proteins to be significantly different between males and females, including Anti-mullerian hormone and multiple insulin growth factors [26].

We have previously shown that the total score of the SDQ is associated with changes in the plasma proteome in adolescents [27]. In another study we also showed association of plasma proteins with the p-factor [28]. These papers show plasma proteins to be a legitimate way of investigating questionnaire-based mental wellness scores. Adjusting for sex in modelling is a routine method, based on the assumption of sex specific plasma differences, however, the influence of sex in these associations have not been previously directly addressed. Here we aimed to systematically investigate the sex-specific associations between plasma protein abundances and the SDQ scores in adolescents. By employing advanced proteomic techniques and bioinformatic analyses, we identified several plasma proteins that were differentially expressed between female and male adolescents. We also found proteins that associated with the SDQ score, and its externalizing of internalizing subscales exclusively in either males or females.

## Methods

### Participant recruitment and sample collection

The studies were reviewed and approved by CEIC Parc Salut Mar Clinical Research Ethics Committee (approval numbers: 2015/6026 Walnuts and 2020/9688–Equal-life). Written informed consent to participate in the original WALNUTs study was provided by the participants’ legal guardian/next of kin. No additional consent was needed for this study, all the participants were offered free tickets to the science museum of Barcelona. The specifics of the Walnuts cohort formation were described in previous publications [29, 30]. The current manuscript used a subset of 372 baseline blood samples before any dietary intervention originally described in a previous work [30]. For this study, a cross-sectional sub-group of 197 samples was used to perform the proteomics analysis. Samples were drawn by a nurse using K2EDTA plus tubes, rested for one hour then centrifuged at 2500 x g for 20 min at 20°C, refrigerated at 4°C, and frozen to −80°C within 4 h after extraction[30], stored at –80°C, and were not thawed until the protein depletion was performed prior to the proteomics analysis. The self-reported SDQ scores were used to assess the psychosocial status reflecting the risk of mental health issues (Table 1).

**Table 1:**
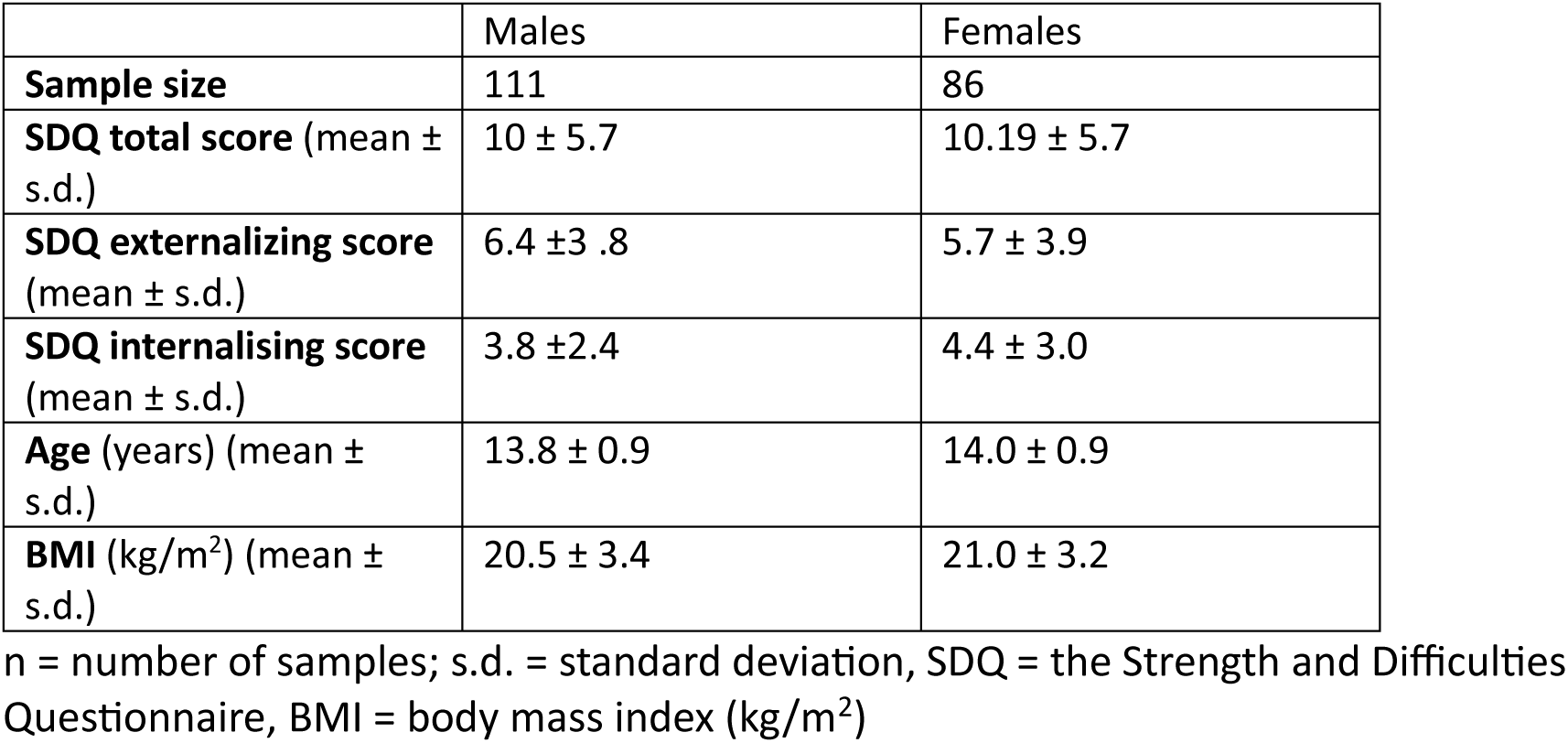
Demographic information of the subjects in the study.

|  | Males | Females |
| --- | --- | --- |
| **Sample size** | 111 | 86 |
| **SDQ total score** (mean ± s.d.) | 10 ± 5.7 | 10.19 ± 5.7 |
| **SDQ externalizing score** (mean ± s.d.) | 6.4 ± 3.8 | 5.7 ± 3.9 |
| **SDQ internalising score** (mean ± s.d.) | 3.8 ± 2.4 | 4.4 ± 3.0 |
| **Age** (years) (mean ± s.d.) | 13.8 ± 0.9 | 14.0 ± 0.9 |
| **BMI** (kg/m<sup>2</sup>) (mean ± s.d.) | 20.5 ± 3.4 | 21.0 ± 3.2 |

n = number of samples; s.d. = standard deviation, SDQ = the Strength and Difficulties Questionnaire, BMI = body mass index (kg/m<sup>2</sup>)

### Proteome analysis

The specifics of the analysis of the first 91 samples are presented in [28]. The analysis specifics for the second part of the cohort were as follows. The second analysis batch contained 120 samples, 106 of them new, and 14 previously analysed samples.

Albumin and IgG represent more than 70% of total protein levels in human plasma samples [31]. Therefore, the depletion of high-abundant proteins is essential to the identification and analysis of low-abundant proteins. The plasma samples were depleted from highly abundant plasma proteins with Thermo Scientific Top14 Abundant Protein Depletion Resin according to manufacturer instructions. The proteins of depleted samples were acetone precipitated. Precipitated proteins were dissolved in 8 M urea in 50 mM Tris-HCl, pH 8. Samples were reduced with 10 mM D,L-dithiothreitol and alkylated with 40 mM iodoacetamide. Samples were digested overnight with sequencing grade modified trypsin (Promega). After digestion peptide samples were desalted with a Sep-Pak tC18 96-well plate (Waters) and evaporated to dryness. Samples were dissolved in 0.1% formic acid and peptide concentration was determined with a NanoDrop device. For DIA analysis 800 ng of peptides were injected and analysed in random order. Wash runs were submitted between each sample to reduce potential carry-over of peptides. 1.3 LC-ESI-MS/MS Analysis The LC-ESI-MS/MS analysis was performed on a nanoflow HPLC system (Easy-nLC1200, Thermo Fisher Scientific) coupled to the Orbitrap Exploris 480 mass spectrometer (Thermo Fisher Scientific, Bremen, Germany) equipped with a nano-electrospray ionisation source and FAIMS interface. Compensation voltages of −40 V and −60 V were used. Peptides were first loaded on a trapping column and subsequently separated inline on a 15 cm C18 column (75 μm x 15 cm, ReproSil-Pur 3 μm 120 Å C18-AQ, Dr. Maisch HPLC GmbH, Ammerbuch-Entringen, Germany). The mobile phase consisted of water with 0.1% formic acid (solvent A) or acetonitrile/water (80:20 (v/v)) with 0.1% formic acid (solvent B). A 120 min gradient was used to elute peptides (62 min from 5 % to 21 % solvent B followed by 48 min from 21 % to 36 % solvent B and in 5 min from 36% to 100% of solvent B, followed by 5 min wash stage with solvent B). Samples were analysed by a data independent acquisition (DIA) LC-MS/MS method. MS data was acquired automatically by using Thermo Xcalibur 4.6 software (Thermo Fisher Scientific). In a DIA method a duty cycle contained one full scan (400 −1000 m/z) and 30 DIA MS/MS scans covering the mass range 400 −1000 with variable width isolation windows.

### Protein identification and quantification analysis

Data analysis consisted of protein identifications and label free quantifications of protein abundances. Data was analysed by Spectronaut software (Biognosys; version 18.0.2). The direct DIA approach was used to identify proteins and label-free quantifications were performed with the MaxLFQ algorithm in Spectronaut. Main data analysis parameters in Spectronaut were: (i) Enzyme: Trypsin/P; (ii) up to 2 missed cleavages (iii) Fixed modifications: Carbamidomethyl (cysteine); (iv) Variable modifications: Acetyl (protein N-terminus) and oxidation (methionine); (v) Precursor FDR Cutoff: 0.01 (vi) Protein FDR Cutoff: 0.01 (vii) Quantification MS level: MS2 (viii) Quantification type: Area under the curve within integration boundaries for each targeted ion (iv) Protein database: Swiss-Prot 2022_05 Homo Sapiens [32], Universal Protein Contaminant database [33]; and (v) Normalisation: Global median normalization. All the peptides were used for quantification.

### Bioinformatic data analysis

To simplify the comparisons, the second analysis batch contained 10 of the same samples, as the first run. The two cohorts were treated as two batches in ComBat batch correction algorithm. The results of the batch correction are presented in Supplementary Figure 1. Data pre-processing and statistical analyses were performed using R (version 4.4.1.). Identified proteins with more than 20% missing values were excluded from the analysis. Sample normalisation was performed using the medianCentering method from the proBatch package [34]. Missing values remaining in the dataset were inputted using the Sample Minimum method, as the most reliable one [35, 36]. Proteins depleted prior to MS analysis that were considered a possible source of bias and thus were excluded.

For the differential abundance analysis limma (v. 3.54.2) [37] package was used, with the SDQ score utilised as a continuous variable. The sex and age of adolescents were included in the linear model to ensure that the proteins reported were associated with SDQ and were not influenced by confounding factors. The differences in protein abundances were expressed as Log2 fold change (ratio of the means of raised (numerator) and low (denominator) SDQ groups). The p-values were adjusted using Benjamini-Hochberg (BH) procedure.

Plasma proteomic datasets from adolescents represent a very low number of all the proteomic datasets [31], so to better investigate the functional enrichment the full list of all proteins found in this study was used as the background gene list in the enrichment analyses. To characterise the enriched pathways related to the identified proteins, significantly differently abundant proteins (adj. p ≤ 0.05) were used in further bioinformatic data analyses. The p-value was adjusted using the BH method. The STRINGdb package was used to retrieve the protein-protein interaction information for the significantly differentially abundant proteins from the STRING database (v. 12) [38] and perform enrichment analysis [39]. Result visualisations were performed using ggplot2 (v3.4.0) [40] package.

## Results

### Protein identification

Mass spectrometry-based proteomics was used to successfully identify 2803 proteins in the WALNUTs plasma samples (mean of 1414 proteins per sample, SD=176). The full list of the proteins is presented in Supplementary File 1. Out of these we removed 119 proteins identified as contaminants or a possible source of bias. Only proteins detected in at least 80% of the samples were used for all the subsequent analyses (N = 802).

### Analysis of sex differences in the plasma proteome

For orthogonal partial least squares discriminate analysis (OPLS-DA), all proteins of the subjects were utilised. Male and female subjects with were distinctly separated in the OPLS-DA plot, indicating satisfactory reproducibility (Fig 1). OPLS-DA plot demonstrated the following measures: Q2Y cumulative = 0.494, R2Y cumulative = 0.852. Sex therefore serves as a reliable separating factor between the plasma proteomes of adolescents in the WALNUTs cohort.

**Figure 1.**
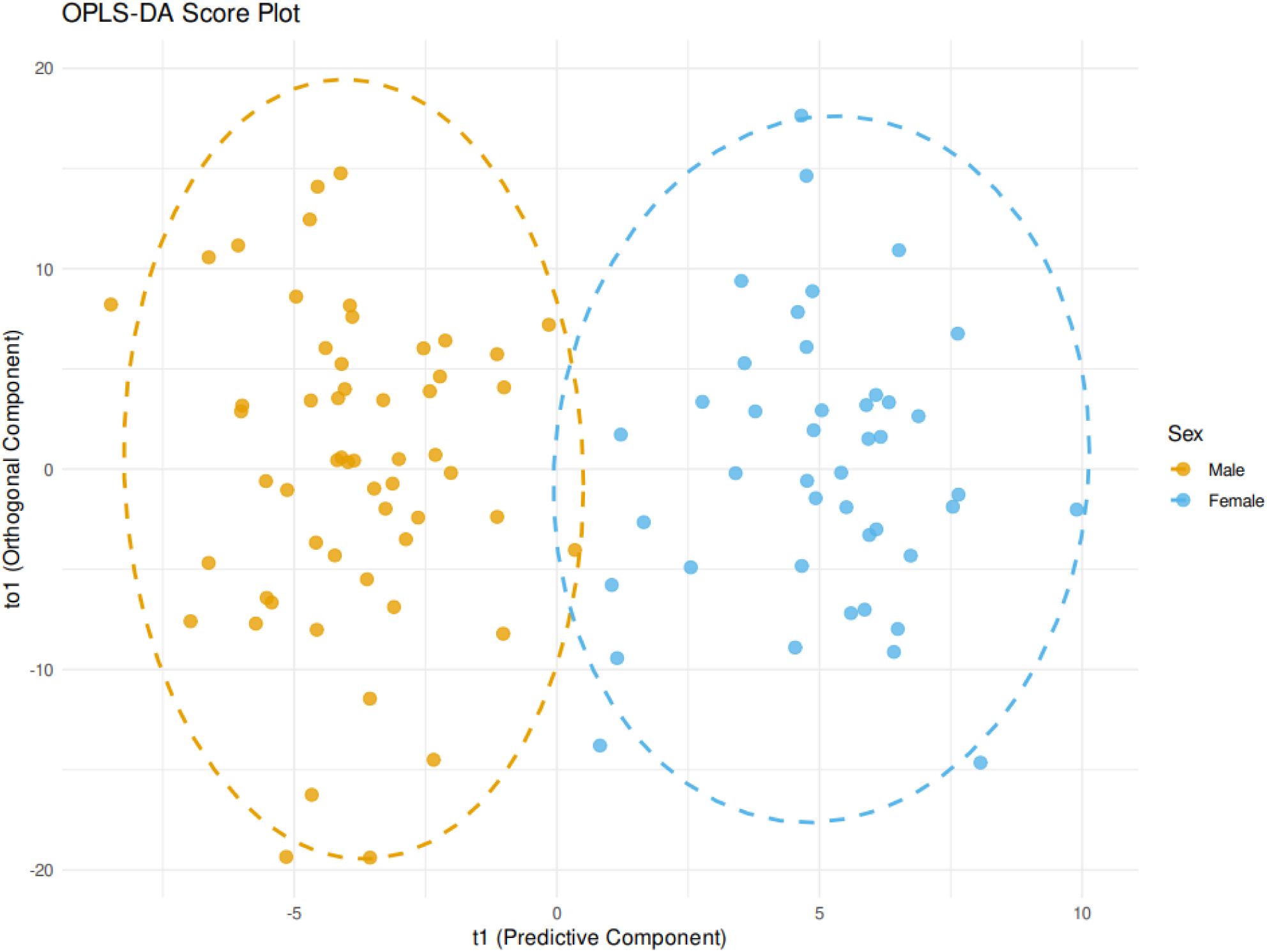
OPLS-DA analysis shows a clear separation of the plasma proteome between the female and male plasma proteomes. Orange colour indicates females and blue indicates males.

To assess the influence of sex on the proteome we used linear modelling with limma. Correcting for age and BMI, linear modelling showed 76 proteins to be different between sexes (Supplementary Table 1). Enrichment analysis for the 76 proteins using Gene Ontology (GO) terms showed that the most significantly enriched categories were cell adhesion, collagen fibril organisation, extracellular matrix organisation, anatomical structure morphogenesis and ossification.

Out of the significant sex-specifically altered plasma proteins (Supplementary Table 1), Vitronectin, Prothrombin, Ficolin 2, and Alpha-1B-Glycoprotein were previously reported to be associated with the SDQ score in adolescents [27]. We therefore split the WALNUTs cohort samples by sex and investigated the sex-specific differences related to the total SDQ score.

### Proteins significantly associated with the full cohort

The full WALNUTs cohort was used to identify the proteins significantly associated with SDQ. We identified 37 proteins to be significantly associated (adjusted p-value < 0.05) with the total SDQ score in this work (Table 3). As splitting the cohort by sex leads to lowered statistical power, we opted to report the nominal p-values for sex-specific analyses (Table 2).

**Table 2.** Proteins associated with SDQ.

| Protein ID | Protein names | Full cohort |  |  | Males |  | Females |  |
| --- | --- | --- | --- | --- | --- | --- | --- | --- |
|  |  | Size effect | P.Value | Adjusted P.Val | Size effect | P.Value | Size effect | P.Value |
| **O15511** | Actin-related protein 2/3 complex subunit 5 | -0.1346 | 0.0013 | 0.0360 | -<br>0.1762 | 0.0047 | - | - |
| **O43895** | Xaa-Pro aminopeptidase 2 | -0.1190 | 0.0022 | 0.0491 | -<br>0.1395 | 0.0112 | - | - |
| **O95445** | Apolipoprotein M | 0.0199 | 0.0009 | 0.0309 | - | - | 0.0292 | 0.0024 |
| **P00450** | Ceruloplasmin | 0.0285 | 0.0001 | 0.0068 | 0.0185 | 0.0379 | 0.0384 | 0.0011 |
| **P00734** | Prothrombin | 0.0242 | 0.0010 | 0.0322 | - | - | 0.0469 | 0.0000* |
| **P01008** | Antithrombin-III | 0.0280 | 0.0000 | 0.0010 | - | - | 0.0439 | 0.0000* |
| **P01833** | Polymeric immunoglobulin receptor | -0.0711 | 0.0000 | 0.0031 | -<br>0.0737 | 0.0009 | -0.0678 | 0.0051 |
| **P02746** | Complement C1q subcomponent subunit B | 0.0210 | 0.0012 | 0.0352 | - | - | 0.0270 | 0.0057 |
| **P02763** | Alpha-1-acid glycoprotein 1 | 0.0352 | 0.0013 | 0.0360 | 0.038 | 0.0103 | - | - |
| **P03951** | Coagulation factor XI | 0.0290 | 0.0002 | 0.0122 | - | - | 0.0421 | 0.0008 |
| **P04003** | C4b-binding protein alpha chain | 0.0278 | 0.0002 | 0.0122 | 0.0205 | 0.0351 | 0.0341 | 0.0039 |
| **P04217** | Alpha-1B-glycoprotein | 0.0202 | 0.0009 | 0.0301 | 0.0178 | 0.0381 | 0.0231 | 0.0105 |
| **P05154** | Plasma serine protease inhibitor | 0.0274 | 0.0020 | 0.0467 | - | - | 0.0442 | 0.0018 |
| **P05155** | Plasma protease C1 inhibitor | 0.0270 | 0.0000 | 0.0051 | 0.018 | 0.0411 | 0.0360 | 0.0003 |
| **P05156** | Complement factor I | 0.0221 | 0.0022 | 0.0491 | - | - | 0.0276 | 0.0092 |
| **P05160** | Coagulation factor XIII B chain | 0.0234 | 0.0023 | 0.0491 | - | - | 0.0380 | 0.0029 |
| **P08603** | Complement factor H | 0.0249 | 0.0001 | 0.0107 | - | - | 0.0370 | 0.0003 |
| **P09603** | Macrophage colony-stimulating factor 1 | -0.1231 | 0.0004 | 0.0189 | -<br>0.1217 | 0.0133 | -0.1335 | 0.0078 |
| **P12955** | Xaa-Pro dipeptidase | 0.0292 | 0.0006 | 0.0228 | - | - | 0.0422 | 0.0060 |
| **P20851** | C4b-binding protein beta chain | 0.0296 | 0.0002 | 0.0122 | - | - | 0.0422 | 0.0015 |
| **P20908** | Collagen alpha-1 | -0.1074 | 0.0001 | 0.0107 | -<br>0.0865 | 0.0150 | -0.1282 | 0.0045 |
| **P31150** | Rab GDP dissociation inhibitor alpha | -0.0965 | 0.0017 | 0.0421 | -<br>0.1251 | 0.0114 | - | - |
| **P35542** | Serum amyloid A-4 protein | 0.0222 | 0.0006 | 0.0228 | - | - | 0.0274 | 0.0067 |
| **P43251** | Biotinidase | 0.0300 | 0.0001 | 0.0113 | 0.0231 | 0.0274 | 0.0378 | 0.0024 |
| **P46531** | Neurogenic locus notch homolog protein 1 | -0.0299 | 0.0010 | 0.0322 | -<br>0.0394 | 0.0032 | - | - |
| **P51693** | Amyloid beta precursor like protein 1 | -0.0736 | 0.0005 | 0.0211 | -0.084 | 0.0028 | - | - |
| **P55287** | Cadherin-11 | -0.1232 | 0.0000 | 0.0051 | -<br>0.1224 | 0.0053 | -0.1219 | 0.0025 |
| **P55899** | IgG receptor FcRn large subunit p51 | -0.1341 | 0.0014 | 0.0360 | -<br>0.1712 | 0.0043 | - | - |
| **P69891** | Hemoglobin subunit gamma-1 | -0.1559 | 0.0012 | 0.0352 | -<br>0.1598 | 0.0245 | -0.1532 | 0.0168 |
| **Q04760** | Lactoylglutathione lyase | -0.1422 | 0.0002 | 0.0122 | -<br>0.1254 | 0.0168 | -0.1589 | 0.0061 |
| **Q15485** | Ficolin-2 | 0.0352 | 0.0003 | 0.0133 | 0.0393 | 0.0019 | 0.0315 | 0.0409 |
| **Q16627** | C-C motif chemokine 14 | -0.0936 | 0.0002 | 0.0122 | -<br>0.0994 | 0.0045 | -0.0858 | 0.0222 |
| **Q6ZP82** | Coiled-coil domain-containing protein 141 | -0.1692 | 0.0000 | 0.0006 | -<br>0.1804 | 0.0003 | -0.1386 | 0.0025 |
| **Q86SF2** | N-acetylgalactosaminyltransferase 7 | -0.1210 | 0.0000 | 0.0035 | -<br>0.1553 | 0.0001* | -0.0884 | 0.0333 |
| **Q9H1U4** | Multiple epidermal growth factor-like domains protein 9 | -0.0414 | 0.0005 | 0.0211 | -<br>0.0569 | 0.0009 | - | - |
| **Q9NSC7** | Alpha-N-acetylgalactosaminide alpha-2,6-sialyltransferase 1 | -0.1349 | 0.0003 | 0.0147 | -<br>0.1649 | 0.0015 | - | - |
| **Q9Y696** | Chloride intracellular channel protein 4 | -0.1352 | 0.0017 | 0.0424 | -<br>0.1695 | 0.0074 | - | - |

The proteins significant with BH correction are denoted with \*

Comparing the list of significant proteins to the sex-specific lists, we see that 15 of the 37 proteins are associated with the full SDQ score in both sexes, 10 are associated only in males, and 12 only in females. Interestingly, only 4 proteins were nominally associated with SDQ in the split analysis, but not in the full cohort (Figure 2).

**Figure 2.**
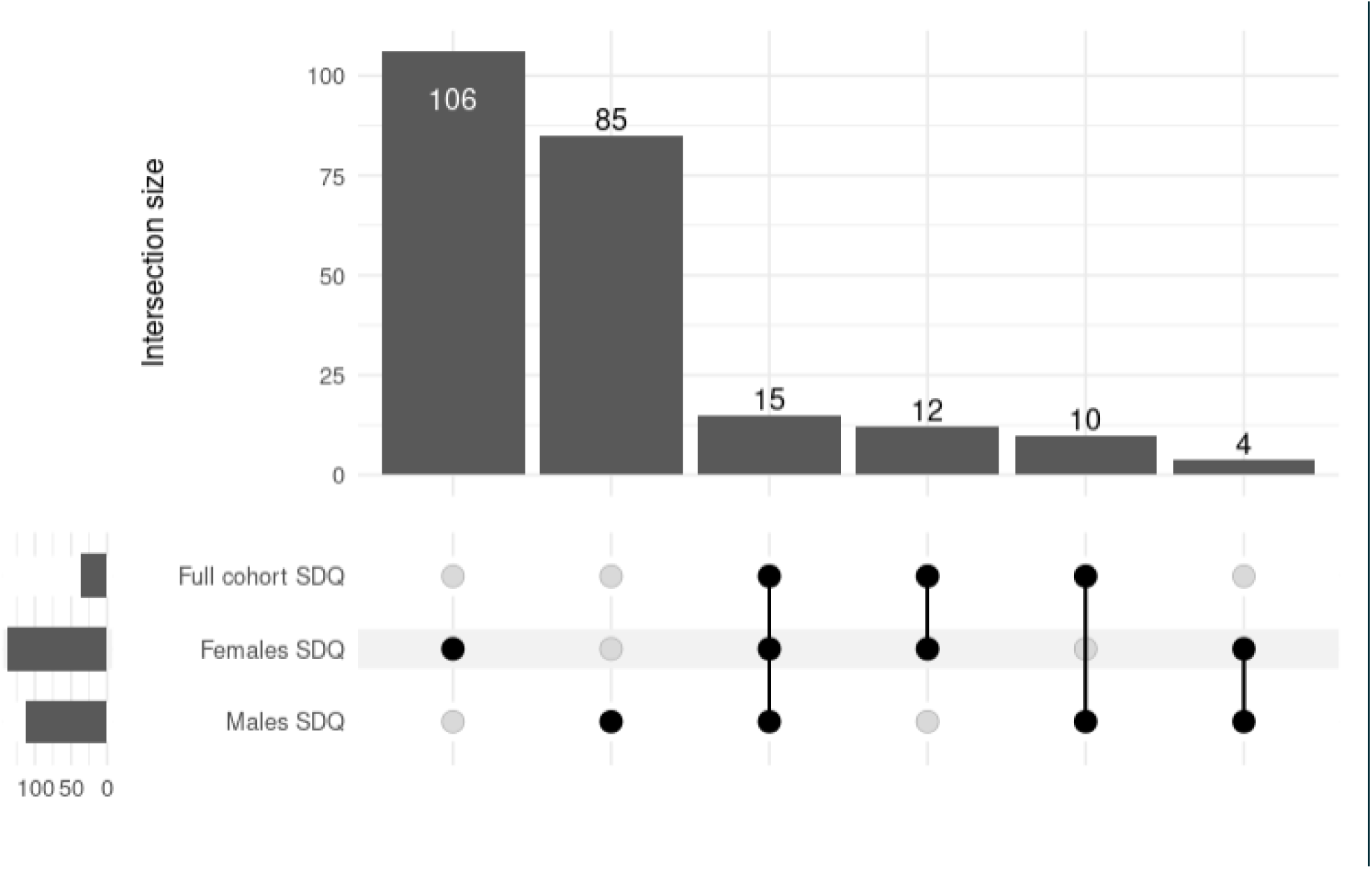
UpSet plot showing the intersections between the set of protein associated with SDQ in the full cohort and in two subsets.

### Protein-protein interactions and enrichment analysis

To predict protein-protein interactions, we next carried out STRING analyses on the 37 DEPs associated with the total SDQ score in the full WALNUTs cohort. The interaction map for the entire cohort, and females and males separately are presented in Figure 3.

**Figure 3.**
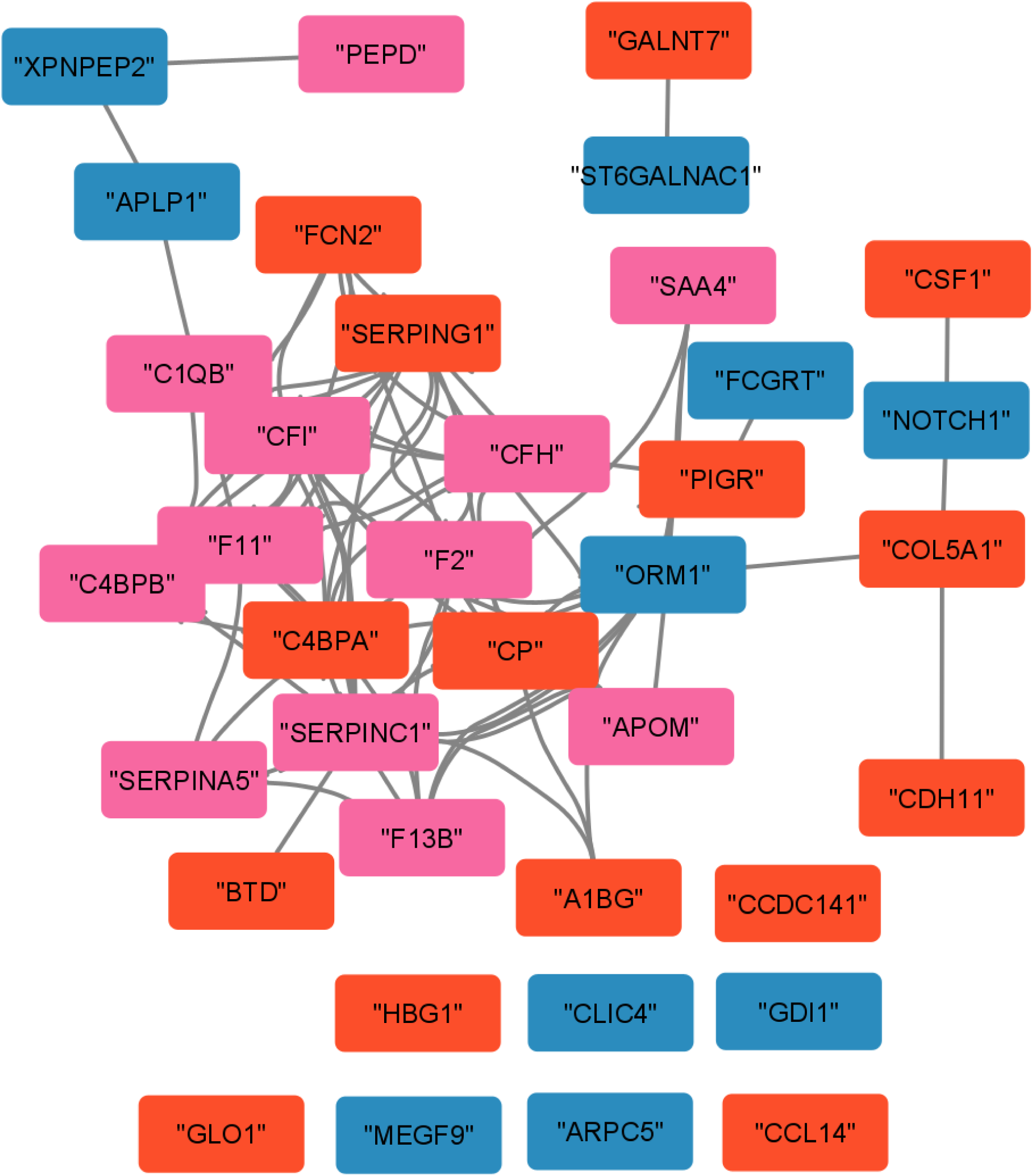
The STRING interaction networks of the proteins, significantly associated with the total SDQ score in the full WALNUTs cohort. The red proteins are those changed in both females and males. Pink boxes denote proteins that are specifically altered only in females and blue denote proteins specifically altered only in males.

We performed enrichment analysis on the three groups indicated in the Figure 3. Only proteins characteristic to the female subset showed significant enrichment the enriched categories included blood microparticles and complement and coagulation cascade. No significant enrichment was found for the other two groups.

### Analysis of sex differences in the plasma proteome associated with the externalizing and internalizing subscales of SDQ

To account for the SDQ subscales, we next analysed the plasma proteomic data separately for females and males in relation to the SDQ internalizing and externalizing subscales. The results of the analysis are presented in the Supplementary File 1. Figure 4 shows the UpSet plot of all the analyses of associations between the total SDQ scores, the externalizing and internalizing subscales of the SDQ, and the protein abundances.

**Figure 4.**
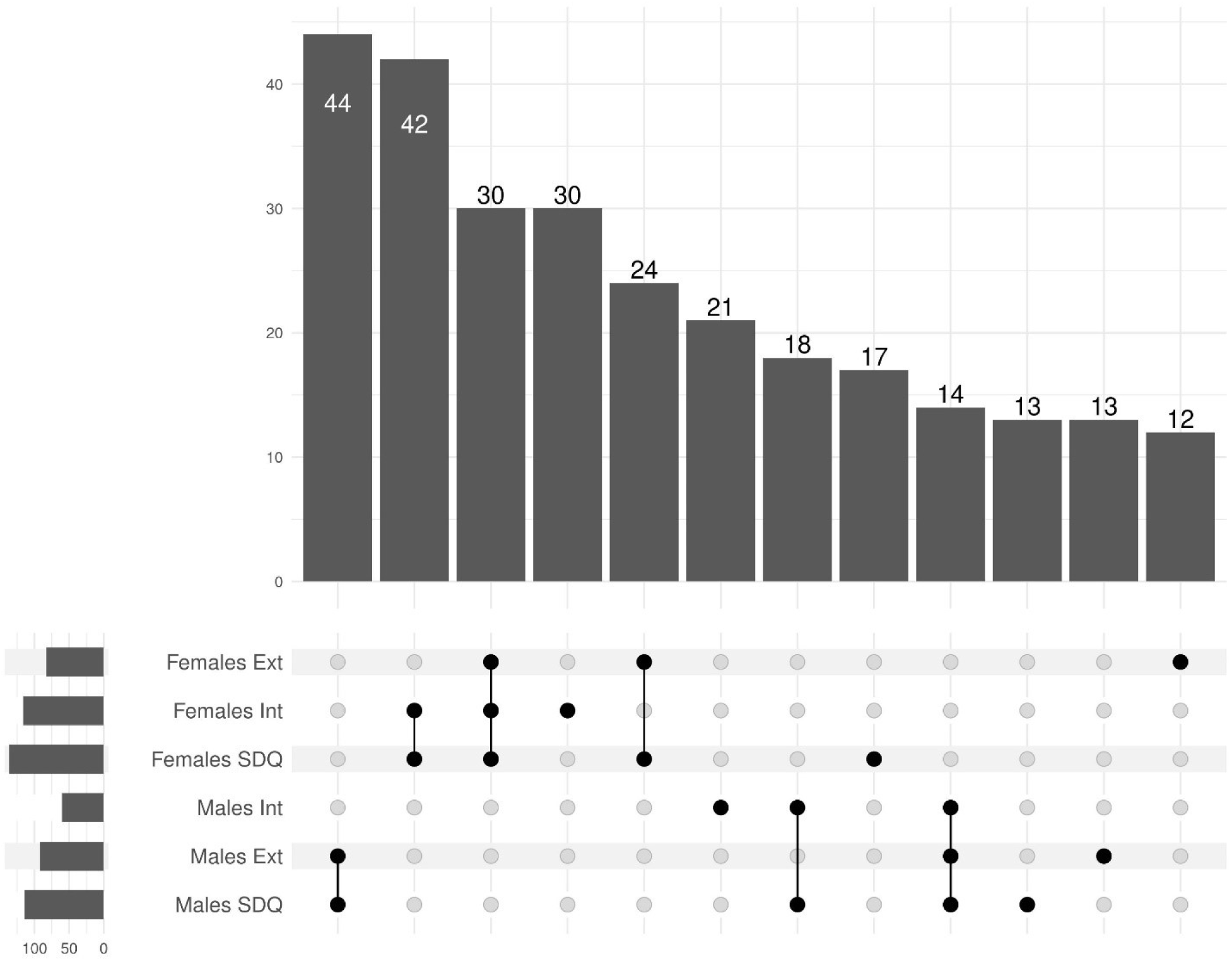
UpSet plot showing the significant proteome features associated with the SDQ score and its sub-scales in females and males. All the intersections with fewer than 10 proteins were removed for readability. The full plot is shown in supplementary figure 2.

The analysis showed that in males, most proteins in association with the total SDQ score were more closely associated with the SDQ externalising scores; 44 proteins were in common between these scores. On the contrary, in females, most proteins in association with the total SDQ score were more closely associated with the SDQ internalizing scores, with 42 common proteins. As the full SDQ is comprised of subscales, the clustering of significantly associated proteins was as expected. The enrichment analysis showed that for females, the proteins were significantly enriched in blood coagulation and complement cascade proteins, whereas no significant enrichment was observed for the male subset.

## 5. Discussion

In this study, we used untargeted LC-MS/MS plasma proteomics to explore the sex-specific associations between plasma protein abundances and mental health indicator scores in adolescents. Our findings provide insights into the proteomic differences of sex-specific psychological and behavioural traits during adolescence, revealing distinct and shared pathways linked to the Strengths and Difficulties Questionnaire (SDQ) scores.

In this study, we used untargeted LC-MS/MS plasma proteomics to investigate the sex-specific associations between plasma protein abundances and mental health indicator scores in adolescents. Overall, 37 proteins were significantly associated with the total SDQ score using the 197 adolescents participating in the WALNUTs cohort. Of all the 76 plasma proteins found to be associated with sex, there were four proteins (Vitronectin, Prothrombin, Ficolin 2, And Alpha-1B-Glycoprotein) that was shown to be associated with the SDQ in the sub-cohort [27]. This coupled with the knowledge that SDQ is sex-specific, motivated us to split the cohort by sex and investigate the associations of the plasma proteome with SDQ separately for females and males.

As splitting the cohort by sex inevitably decreases the power of the statistical analysis we opted to use nominal p-values for these analyses. The analyses of the split cohorts showed individual proteins to be significantly and sex-specifically associated with the total SDQ score; N-acetylgalactosaminyltransferase 7 in males, and prothrombin and antithrombin-III in females. In a previous study, the level of N-acetylgalactosaminyltransferase in the brain was connected to sex differences in depressive, anxious behaviours in a rat model [41]. The female-specific proteins both play a role in blood coagulation which has been previously linked to depression and anxiety [42], and is in line with our previous study [27]. The deregulation of blood coagulation proteins was previously reported to be associated with acute stress [43] and depression [44].

Our analyses did, however, reveal the levels of IGFBP2 to be significantly higher in males than in females, in line with results previously reported by Liu et al.[26]. Additionally, we found sex-hormone-binding globulin proteins to also be associated with sex, possibly connected to different levels of sex hormones, which is to be expected[45]. We also identified differences in proteins related to bone structure and ossification, which is consistent with the significant skeletal development occurring during puberty [46]. It was also shown, that although males tend to produce more inflammatory responses before puberty, after the puberty, inflammation is higher in females than in males [23], which makes the investigation of inflammation-associated pathways to be complicated in mixed-sex cohorts. Given that puberty also occurs at different ages for males and females, developmental differences can additionally influence protein abundances [47, 48], further emphasising the importance of analysing the sexes separately, even in an age-balanced cohorts like WALNUTs.

The shared proteins between sexes have the same direction of the association and similar size effects. We can assume that the 15 proteins common to both sexes and associated with the SDQ score play a role in the underlying differences between the low and high-SDQ individuals. Of these proteins, amyloid beta precursor like protein 1 (APLP1) and ficolin 2 were previously described as associated with SDQ [27]. When analysing the sex-specific associations we saw that the bulk of the proteins – 106 for females and 85 for males – were different. This suggests that distinct mechanisms underlie the behavioural and psychological traits of each sex in addition to the shared ones. For females, proteins enriched in blood coagulation pathways and complement cascades were prominent, whereas males did not exhibit similar enrichment patterns.

When dissecting SDQ into externalizing and internalizing subscales, we observed that the proteomic associations in males were more closely linked to externalizing sub-score, while in females internalizing sub-score had more proteins in common with the full SDQ score. This aligns with behavioural research suggesting that males often exhibit more outward-directed behavioural challenges, while females are more prone to internalizing conditions such as anxiety and depression [11, 41]. The distinct sex-associated proteomic signatures reinforce the need to consider sex differences when evaluating biological markers for psychological and behavioural traits. The observation that SDQ subscales—externalizing in males and internalizing in females—show distinct proteomic signatures underscores the need for a nuanced approach to studying adolescent psychological development. The involvement of immune response pathways and the enrichment of blood coagulation proteins in females suggest that biological mechanisms underlying psychological traits may vary substantially between sexes. Immunological markers were previously associated with depression and anxiety symptoms, some authors suggesting a direct link between deregulation of immune response to depression [49, 50]. These findings provide a foundation for further investigations into the interplay between proteomic profiles, sex-specific development, and behavioural outcomes.

There exist some limitations in this study. First, the cohort contains only adolescents from a specific region of Spain. Although they are of quite similar age, the range is more than two years, which can be critical when investigating adolescents. In addition, the entire cohort was split into two for sex-specific analyses, which made the number of common proteins low and increased the analysis complexity. The second sequencing batch is homogenous with regards to SDQ, with most participants having low to moderate total SDQ scores, which makes the full cohort not much better at detecting associations with extreme SDQ values. Furthermore, the protein depletion method used in the plasma analyses affects the abundance estimation of multiple plasma proteins - other depletion methods can potentially yield more reliably detected proteins and improve repeatability [51]. Despite the limitations of this study, including a relatively homogeneous cohort and reduced statistical power in sex-specific analyses, the results pave the way for future research. Our findings emphasise the importance of analysing male and female proteomes separately to uncover distinct biological pathways, particularly those related to coagulation, immune responses, and psychological health.

Future investigations should focus on validating the results in independent sets of samples using larger, multi-cohort datasets to improve the reproducibility and usefulness of the found associations. The statistical power of the OMICs studies should also be considered when planning future work with cohorts. To improve the results analysing each sex separately may be a feasible way forward for analysing mental health, specifically in adolescents. Furthermore, integrating longitudinal data could reveal how proteomic profiles evolve during adolescence and their relationship with long-term psychological outcomes.

## Data availability

The data analysed in this study is subject to the following licenses/restrictions: The Walnuts data is not publicly available due to the restrictions of informed consent. The data contains personal information of children and according to the ethical approval, they should be kept confidential. Proteomics data is not publicly available due to the restrictions of informed consent. Reasonable requests to access these datasets should be directed to Katja Kanninen, University of Eastern Finland. To ensure the protection of privacy and compliance with national data protection legislation, a data use/transfer agreement is needed, the content and specific clauses of which will depend on the nature of the requested data.

## Acknowledgements

We gratefully acknowledge the contribution of the Early Environmental quality and life-course mental health effects (Equal-Life) project team, Ruth Koole and Rik Bogers (RIVM) in providing the selection protocol, the Turku Proteomics Facility team supported by Biocenter Finland for mass spectrometry, and the laboratory assistance of Ms. Mirka Tikkanen.

This WALNUTs project was supported by Instituto de Salud Carlos III through the projects ‘CP14/00108, PI16/00261, and PI21/00266’ (co-founded by the European Regional Development Fund ‘A way to make Europe’). JJ holds a Miguel Servet-II contract (grant CPII19/00015) awarded by the Instituto de Salud Carlos III (confounded by the European Social Fund “Investing in your future”). The California Walnut Commission (CWC) gave support by supplying the walnuts for free for the Walnuts Smart Snack Dietary Intervention Trial. The funders had no role in the study design, collection, management, analysis, and interpretation of data, the writing of the report, or the decision to submit it for publication.

## Author Contributions

KMK, ISM, and IvK designed the study. IvK and JJ guided the selection process and provided the samples. AA performed statistical and bioinformatics analyses, pathway analysis, planning and literature analysis. AA, A-KP and KMK drafted the manuscript. All authors read and approved the final version of the manuscript.

## Supplementary materials

**Supplementary figure 1.**
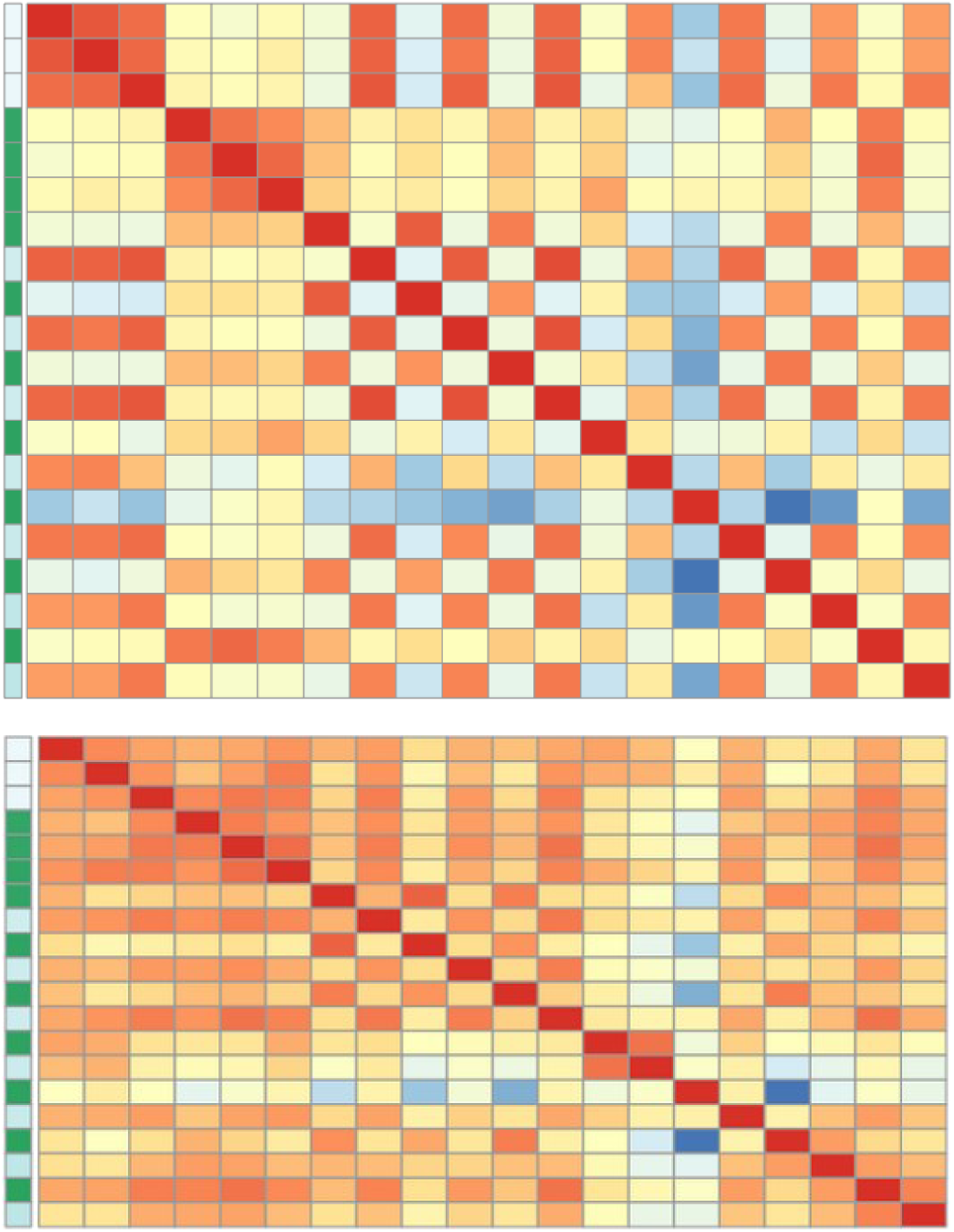
The heatmaps of sample correlations. A - samples before batch correction, B –samples after batch correction. The first 3 samples are from the first sequencing batch, the next 4 are from the second sequencing run, and the rest are repeating samples from the first batch followed by the second batch. Red signifies high correlation; blue signifies low correlation.

**Supplementary figure 2.**
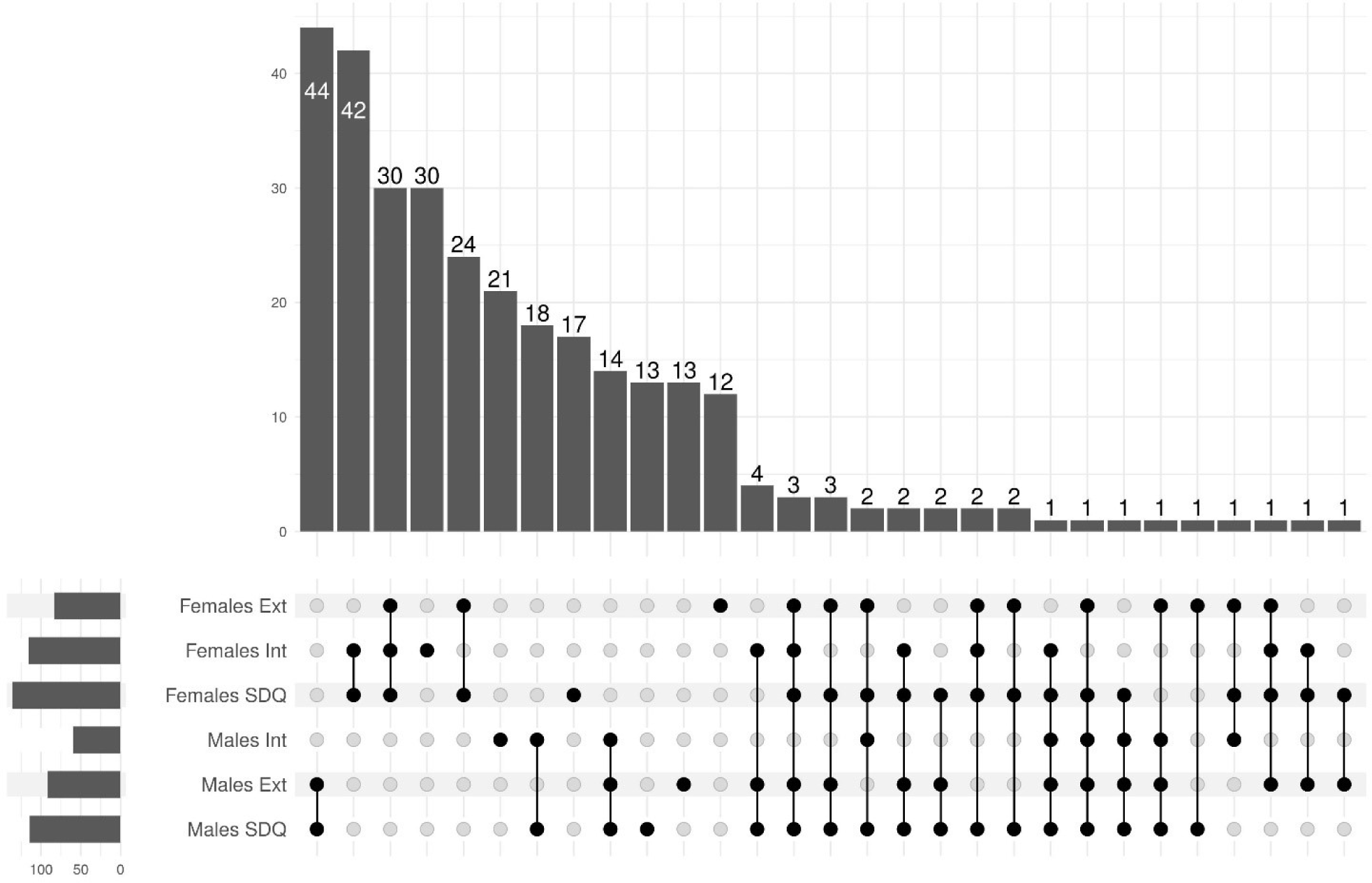
The full UpSet plot showing the significant proteins associated with the SDQ sub-scales in females and males.

**Supplementary table 1.**
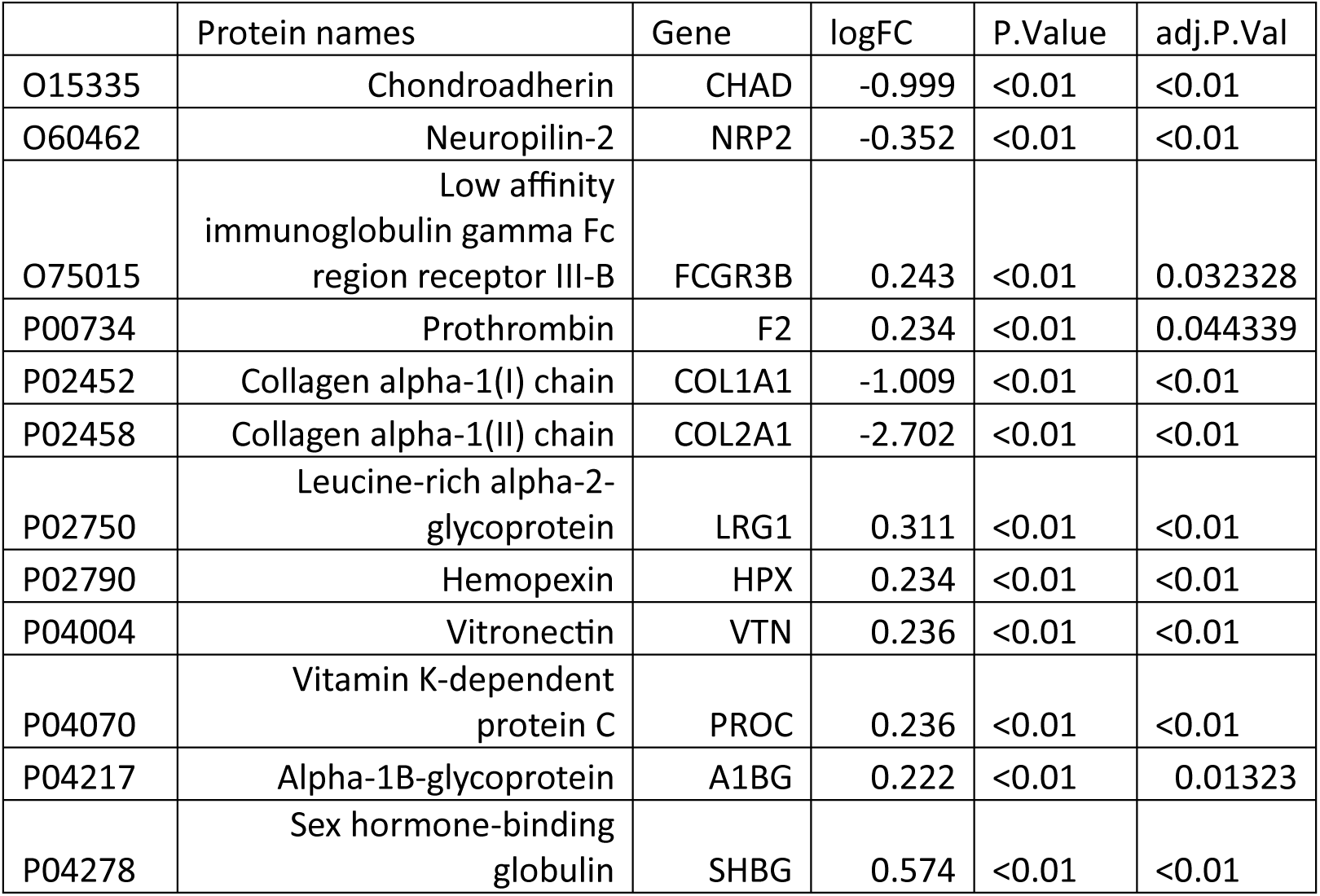
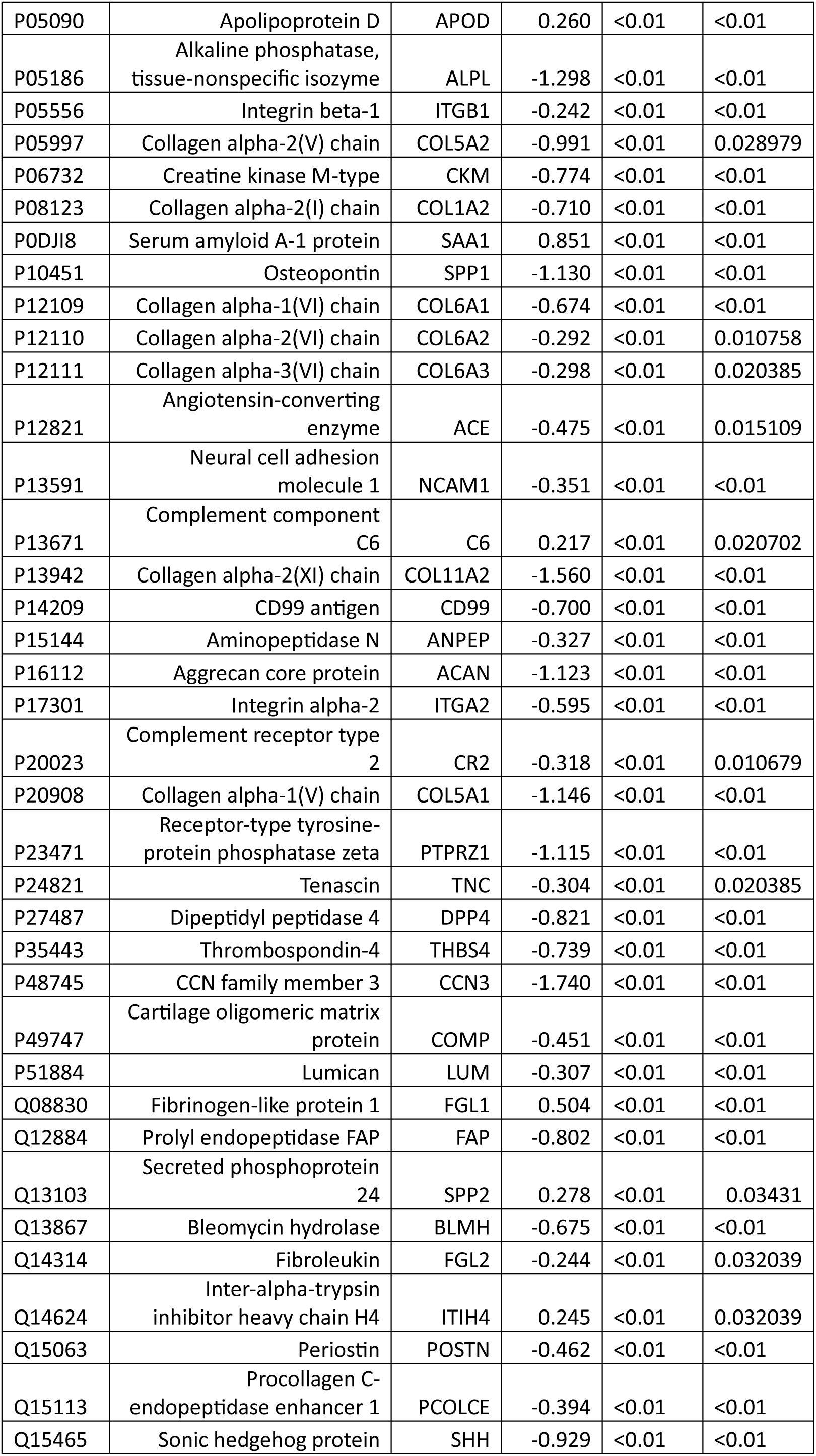
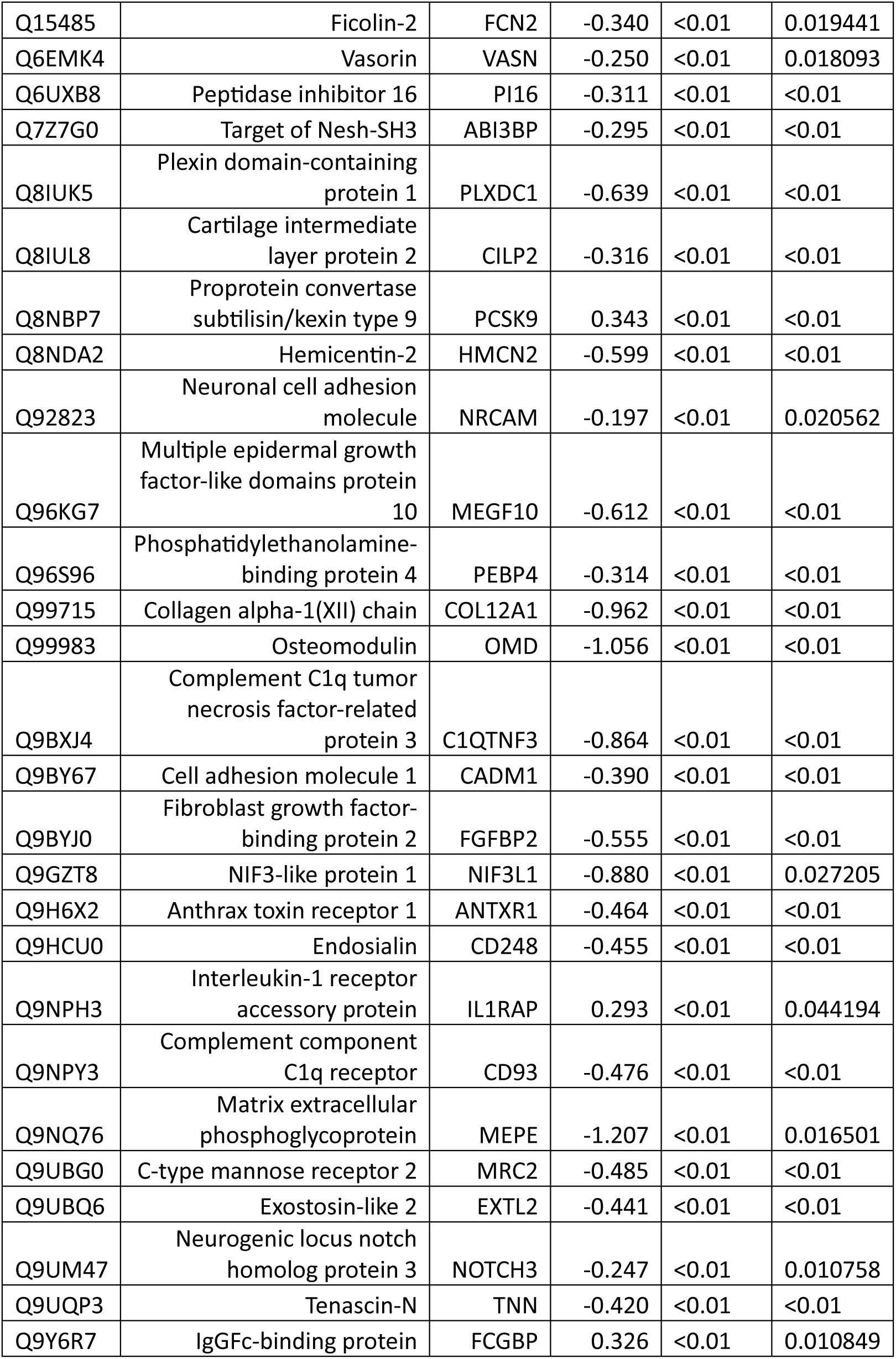
Proteins significantly associated with sex.

